# Dynamic, stage-course protein interaction network using high power CpG sites in Head and Neck Squamous Cell Carcinoma

**DOI:** 10.1101/2021.06.30.21259548

**Authors:** Arsalan Riaz, Maryam Shah, Saad Zaheer, Abdus Salam, Faisal F Khan

## Abstract

Head and neck cancer is the sixth leading cause of cancer across the globe and is significantly more prevalent in South Asian countries, including Pakistan. Prediction of pathological stages of cancer can play a pivotal role in early diagnosis and personalized medicine. This project ventures into the prediction of different stages of head and neck squamous cell carcinoma (HNSCC) using prioritized DNA methylation patterns. DNA methylation profiles for each HNSCC stage (stage-I-IV) were used to extensively analyze 485,577 methylation CpG sites and prioritize them on the basis of the highest predictive power using a wrapper-based feature selection method, along with different classification models. We identified 68 high-power methylation sites which predicted the pathological stage of HNSCC samples with 90.62 % accuracy using a Random Forest classifier. We set out to construct a protein-protein interaction network for the proteins encoded by the 67 genes associated with these sites to study its network topology and also undertook enrichment analysis of nodes in their immediate neighborhood for GO and KEGG Pathway annotations which revealed their role in cancer-related pathways, cell differentiation, signal transduction, metabolic and biosynthetic processes. With information on the predictive power of each of the 67 genes in each HNSCC stage, we unveil a dynamic stage-course network for HNSCC. We also intend to further study these genes in light of functional datasets from CRISPR, RNAi, drug screens for their putative role in HNSCC initiation and progression.

## Introduction

Head and neck cancer is the sixth leading cause of cancer across the globe and is more prevalent in the developing countries of South Asia, including Pakistan [2]. Cancer of the head and neck region is often diagnosed at advanced stages leading to a reduced survival rate of 20% with significant mortality and morbidity [3]. The progression of the cancer is classified using the staging system of the American Joint Committee on Cancer (AJCC) to determine the treatment for the patient [4]. However, the outcomes of the treatment depends upon the molecular biology of the cancer cells and it is important to identify and understand the actionable molecular signature for better and personalized treatment at any particular stage of the cancer [5].

Epigenetic modifications are emerging molecular features that are stable heritable phenotypes resulting from changes in a gene without alterations in the DNA sequence [6]. The most studied epigenetic modification of genes is DNA methylation, which can alter the gene expression without altering the DNA sequence, preferably in the CpG islands. CpG islands are regions in the genome with a high frequency of CpG sites where the percentage of cytosine and guanine is greater than 50 %. DNA methylation is a process in which a methyl group is attached mostly to a cytosine base in DNA and plays an important role in regulating gene expression [7]. Gene expression can be significantly regulated by alterations in DNA methylation patterns and these alterations can result in mutational effects that can lead to cancer [8]. Studies show that DNA methylation markers can be used for the diagnosis of common cancers [9, 10]. Advancements in high-throughput sequencing with platforms such as Illumina Infinium Human Methylation 450K BeadChip array, which can cover over 480K CpG sites and target 96% of CpG islands in the human genome has enabled large studies such as The Cancer Genome Atlas (TCGA) to understand the methylation patterns in different cancer types and their progression [11, 12]. DNA methylation is an important contributor to the pathophysiology of the various stages of cancer making it important to understand the role of specific methylation events in each stage of a particular cancer. If we understand the dynamics of DNA methylation in the specific stages of cancer then it has the potential to give rise to new precision medicine therapies for each patient based on their specific tumor [13].

Recently, Veeramachaneni et al (2019) reported changes in the genomic landscape in the development of head and neck squamous cell carcinoma (HNSCC) from premalignant lesions to malignancy and lymph node metastasis. They showed that known cancer drivers have significantly increased frequency of somatic copy number alterations, maybe due to deletion or promoter hypermethylation, when transitioning from premalignant lesions to HNSCC [14]. In 2013, a study was conducted by Kang et al, to identify those genes which undergo methylation alterations as the tumor progresses from benign to malignant form. For this purpose, genome-wide methylation databases of breast cancer cell lines from stage I to stage IV were analyzed, where they found the promoter methylation level of some genes consistently increased through normal cell lines to stage IV cell lines of breast cancer [15]. Additionally in the same year, methylation patterns of selected genes were analyzed for which previous breast cancer DNA methylation reports were available and they also concluded that the level of aberrant DNA methylation is higher in late-stage compared with early stage of invasive breast cancers [16]. Eventually, such studies encourage data scientists to analyze complex databases and build statistical models for helping in cancer diagnosis and prognosis.

Advancements in modern machine learning and artificial intelligence techniques have aided researchers to detect and predict common types of cancer better and faster than clinicians using complex datasets [17]. Research studies have applied probabilistic methods, and machine learning algorithms to microarray gene expression data for the classification of cancer types [18-20]. In a recent study, a deep neural network was applied to predict the unknown primary origin of the cancer cell with high accuracy using DNA methylation datasets. The study used one-way ANOVA and Turkey’s HSD to prioritize 10360 methylation sites from a total of 27K sites. These sites were used in the input layer of the neural network [21]. However, there is a need for better prioritization of DNA methylation sites to understand the potential role of the genes that harbor these sites in disease onset.

For the prioritization of methylation sites, machine learning feature selection algorithms can be applied to extract the subset of features from the information system that can predict the target outcomes. In 2019, Kaur et al predicted early (stage I) and late (stage II, III, IV) stages of Liver Hepatocellular Carcinoma using differentially expressed transcripts and methylation CpG sites, identified through the Yuen-Welch test and feature selection techniques available in the WEKA software package and Scikit learn package. In their results, they achieved a respectable accuracy of 75.27 % for stage prediction using Random Forest with 21 CpG sites selected from 447 CpG differentially methylated sites [22]. One drawback of selecting CpG sites from differentially methylated sites is to lose those sites with a mid-methylation score, which might have prediction power as well as biological and clinical meaning. A feature selection algorithm, known as the Boruta algorithm, can extract features of significant relevance from an information system regardless of the size of the dataset. The Boruta algorithm was developed by Miron B. Kursa and Witold R. Rudnicki as an R package at the University of Warsaw in 2010 [1]. The Boruta algorithm is a Random Forest based wrapper feature selection method that can provide an unbiased and stable prioritization of significant CpG sites from a total of 485,577 CpG sites. The key idea behind the Boruta algorithm is to make a randomized copy, also called the shadow features, of the information system then merge this copy with the original dataset and build the Random Forest (RF) classifier for this extended information system. After this, the algorithm performs predefined RF iterations and the replicated features are randomized before each iteration. The importance of all the features is computed in each iteration. A feature is considered important for a single iteration if its importance is higher than the maximal importance of all the randomized features and if the importance is below the maximal importance then the feature is removed from the information system. The features can be ranked based on the mean importance which is the mean of the importance computed at each iteration [23].

In our study, we have built three machine learning-based classification models to predict pathological stages (stage 1, stage 2, stage 3, stage 4a) of primary tumors from HNSCC patients based on a set of key methylation sites (CpG sites) identified using the Boruta algorithm, regardless of the primary sites or demographics of the patients [24]. We have also analyzed the feature contribution for each stage in the high-performance classification model to identify sites and their genes which offered a higher prediction power. Further, GO term enrichment analysis, KEGG pathway analysis, PPI network analysis was also performed for the associated genes of the identified CpG sites.

## Results

### The Boruta algorithm identified 68 high-power CpG sites

In phase 1, after intense processing of 13.7 hrs, 294,603 CpG sites were reduced to 1580 important CpG sites. In phase 2, a file of 1580 important CpG sites from phase 1 was prepared, and Boruta was applied with maximum runs of 50,000 iterations and default parameters. Phase 2 provided 68 important CpG sites out of 1580 and was confirmed in phase 3, where no more unimportant CpG sites were found (Figure 1). Time taken by runs in phase 2 and phase 3 was 1.4 hr and 10.7 mins, respectively. These 68 CpG sites are located in 67 genes and are ranked by the mean importance of the Boruta algorithm. CpG sites of SPO11, CMSS1, EFR3B, and ESD were the most important (Table 1).

**Table 1:**
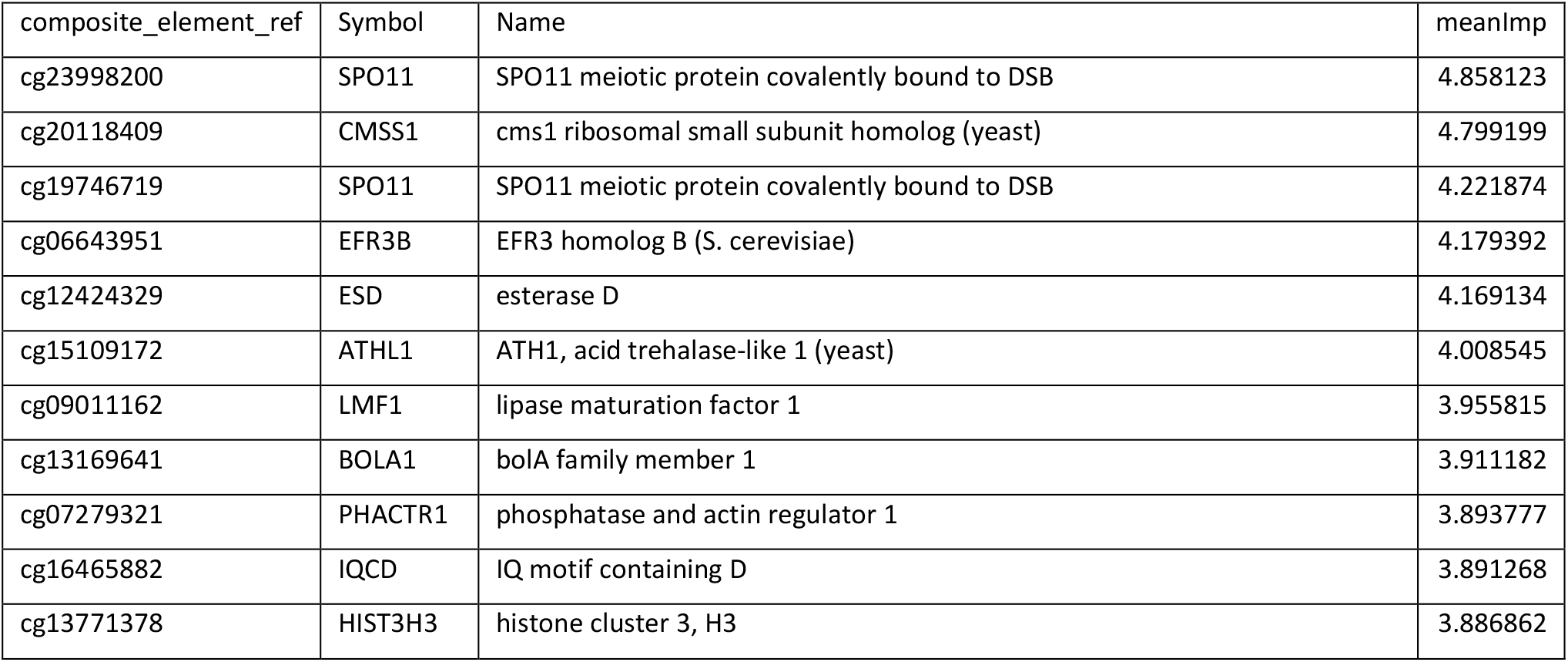
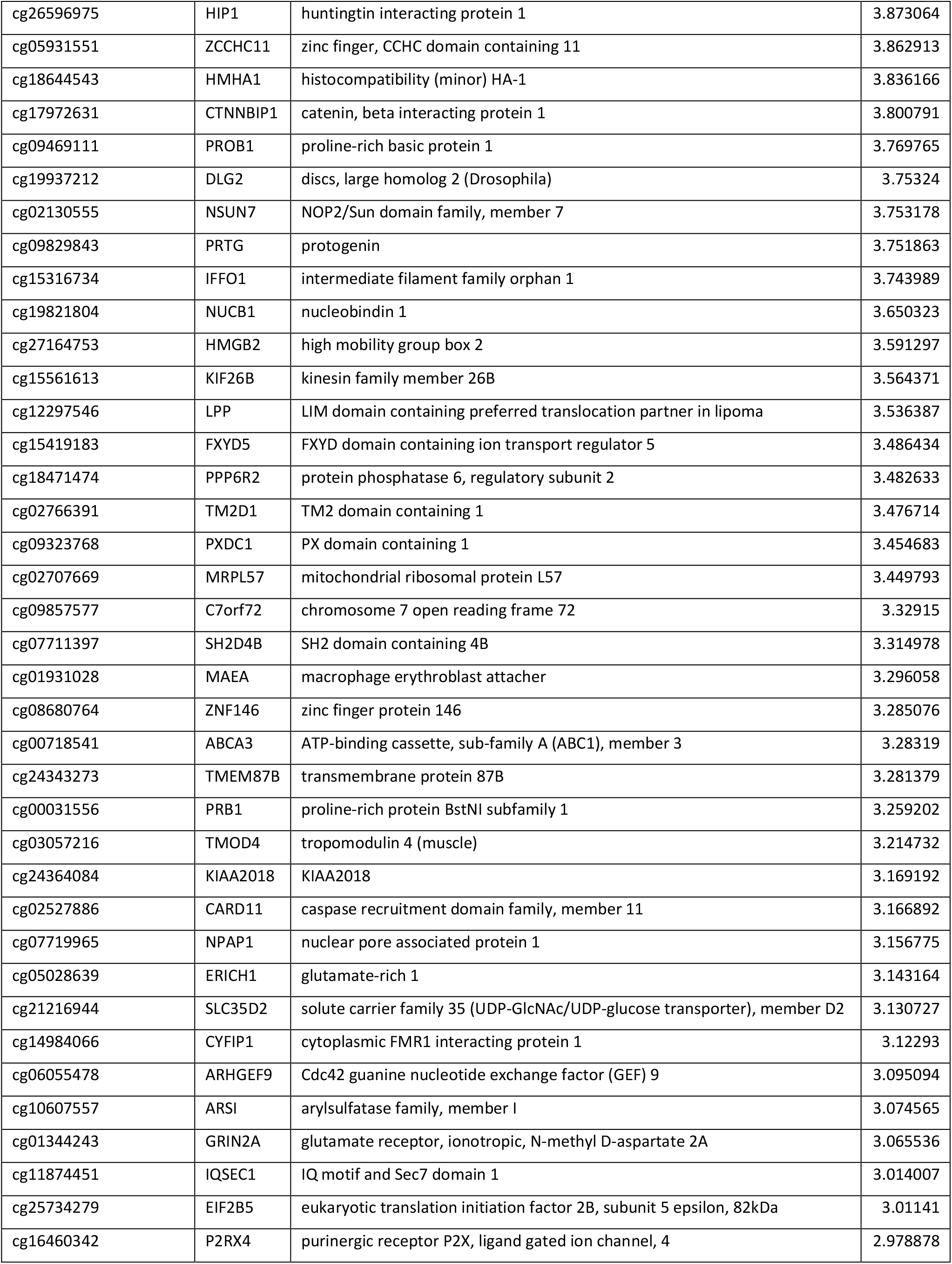
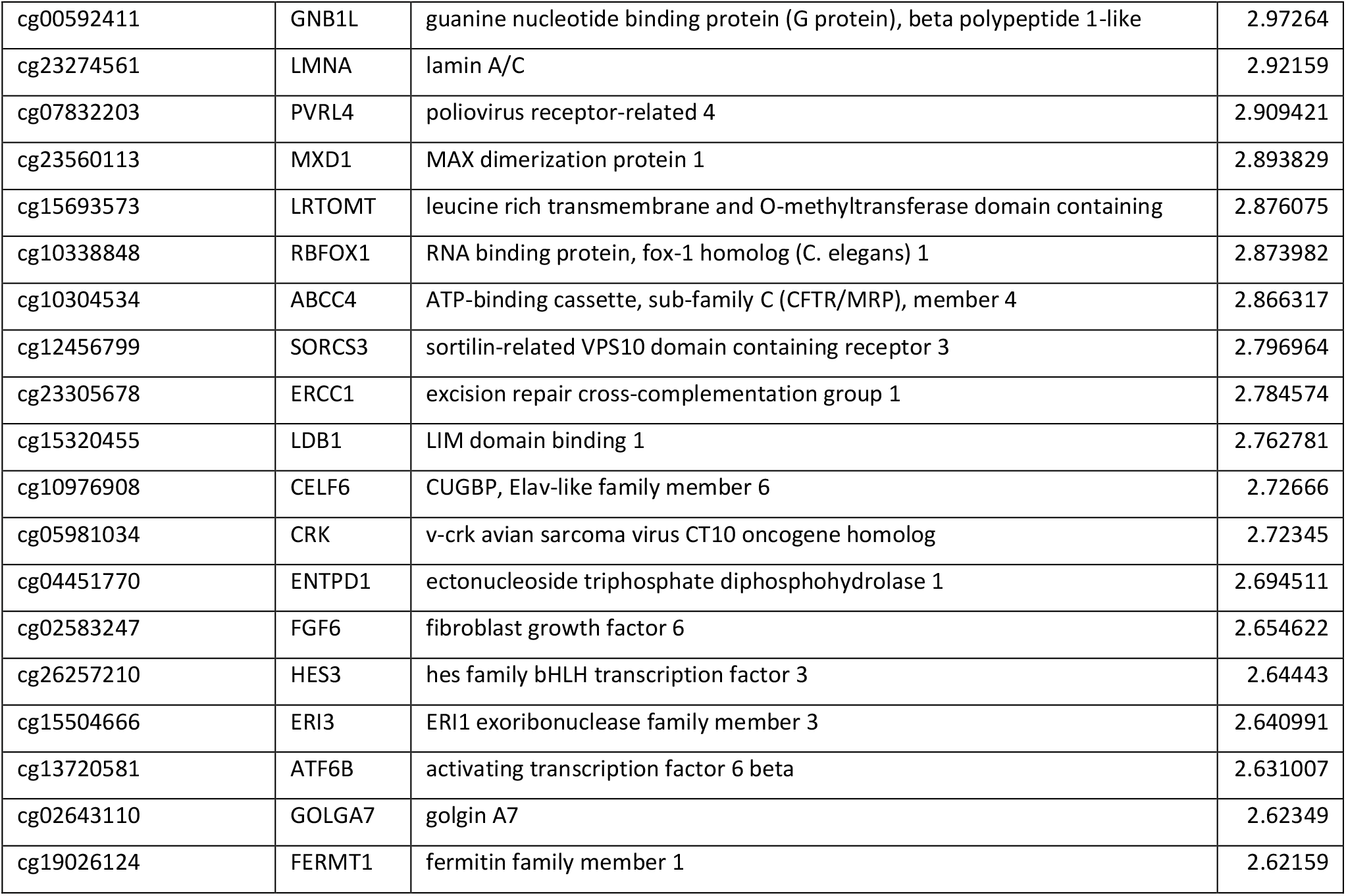
Boruta power predictive CpG sites. Boruta extracted CpG sites with their corresponding Genes and mean importance calculated by the Boruta algorithm.

**Figure 1:**
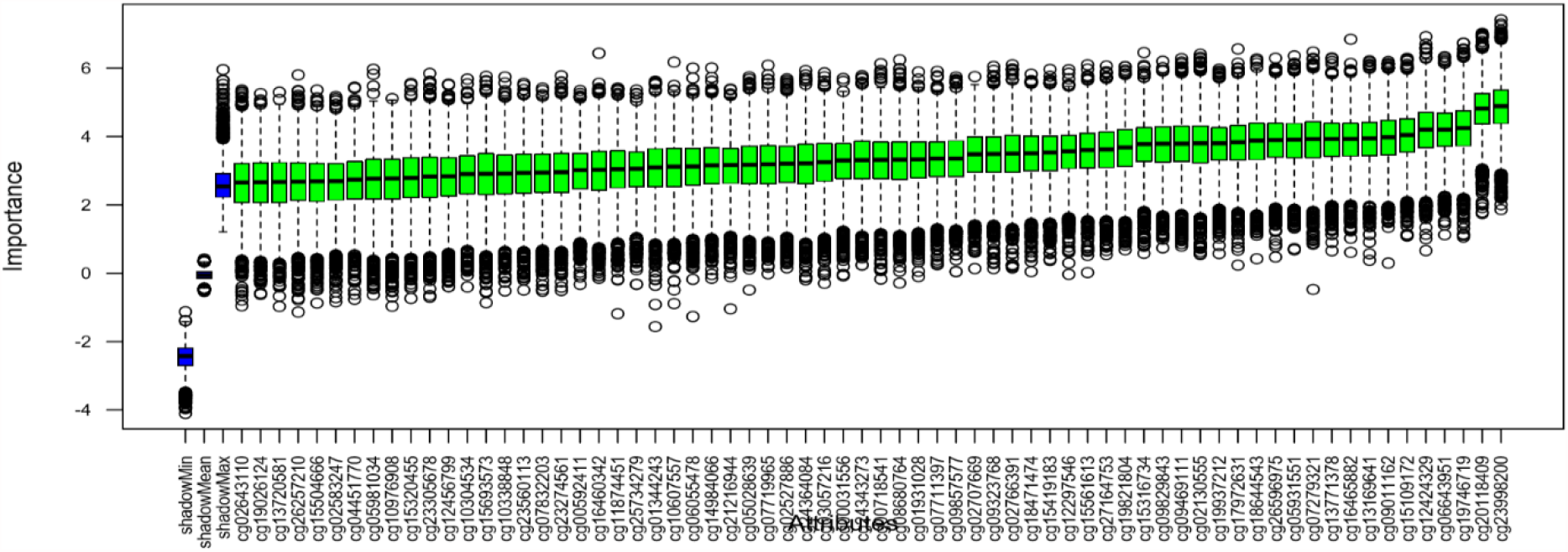
Boruta algorithm box plot. The Boruta algorithm identified 68 CpG sites as high powered which are used as primary features in the prediction models. X-axis represents the composite element REF (CpG sites) and y-axis represents the importance of the sites. Steeper box plot represents high importance composite element REF.

### Random Forest classifier staged primary tumor with 90.62% accuracy

After obtaining our 68 CpG sites, we evaluated three machine learning models i.e. Random Forest (RF), Support Vector Machine (SVM), and K-Nearest Neighbor (KNN) for the classification and prediction of pathological stages (stage 1, stage 2, stage 3, stage 4a) of HNSCC, with 10-fold cross-validation repeated 30 times using the R caret package. Random Forest outperforms the SVM and KNN classifiers in predicting the pathological stages of the testing dataset (30%) after training the machine learning models using the training dataset (70%). Random Forest classifier predicted the stages with an accuracy of 90.62% (95% CI: 74.98-98.02%) with a kappa value of 0.875 and P-value of 7.507e-15, the best in predicting the stages in the test data than SVM and KNN. Where SVM with linear kernel had an accuracy of 68.75% (95% CI: 49.99-83.88%) with a kappa value of 0.583 and a P-value of 2.406e-07. SVM with radial kernel had an accuracy of 71.88% (95% CI: 53.25-86.25%), kappa value of 0.625, and P-value of 3.411e-08. KNN performed with the lowest accuracy of 56.25% (95% CI: 37.66-73.64%), kappa value of 0.4167, and P-value of 1.602e-04 (Figure 2).

**Figure 2:**
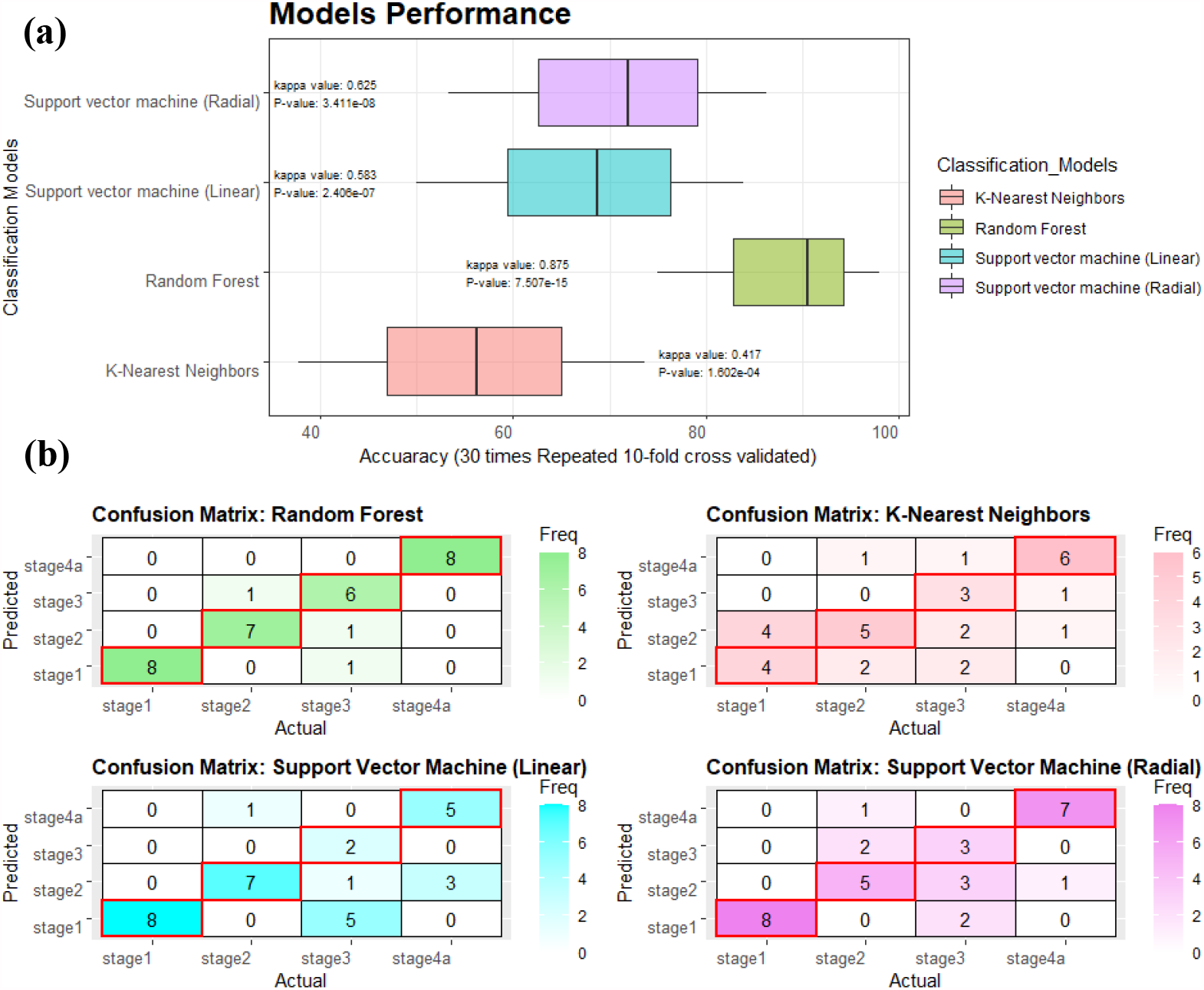
Machine learning models performance. (a) Performance evaluation of machine learning models where boxplots present accuracies with 95% confidence interval along with corresponding Kappa and p-values. (b) Confusion matrices for each machine learning models where cells with red boundries indicates the number of correctly classified samples for each stage.

### Feature contribution analysis

Considering the Random Forest as the best model, we analyzed the feature contribution of CpG sites to each class (pathological stage) using the varImp function of the caret package in R. Feature contribution provides the measure of the influence of features (CpG sites in our case) on the prediction outcome [25], where the outcome is the pathological stage of HNSCC. We observed the contribution of most sites for stage 4a while the least for stage 1, consistent with the fact that tumor progression is attributed to the accumulation of alterations overtime driving the course of the disease [26]. This analysis of feature contribution might be useful in predicting the stage specificity of a gene and also providing a dynamic stage-course view of HNSCC progression (Figure 3).

**Figure 3:**
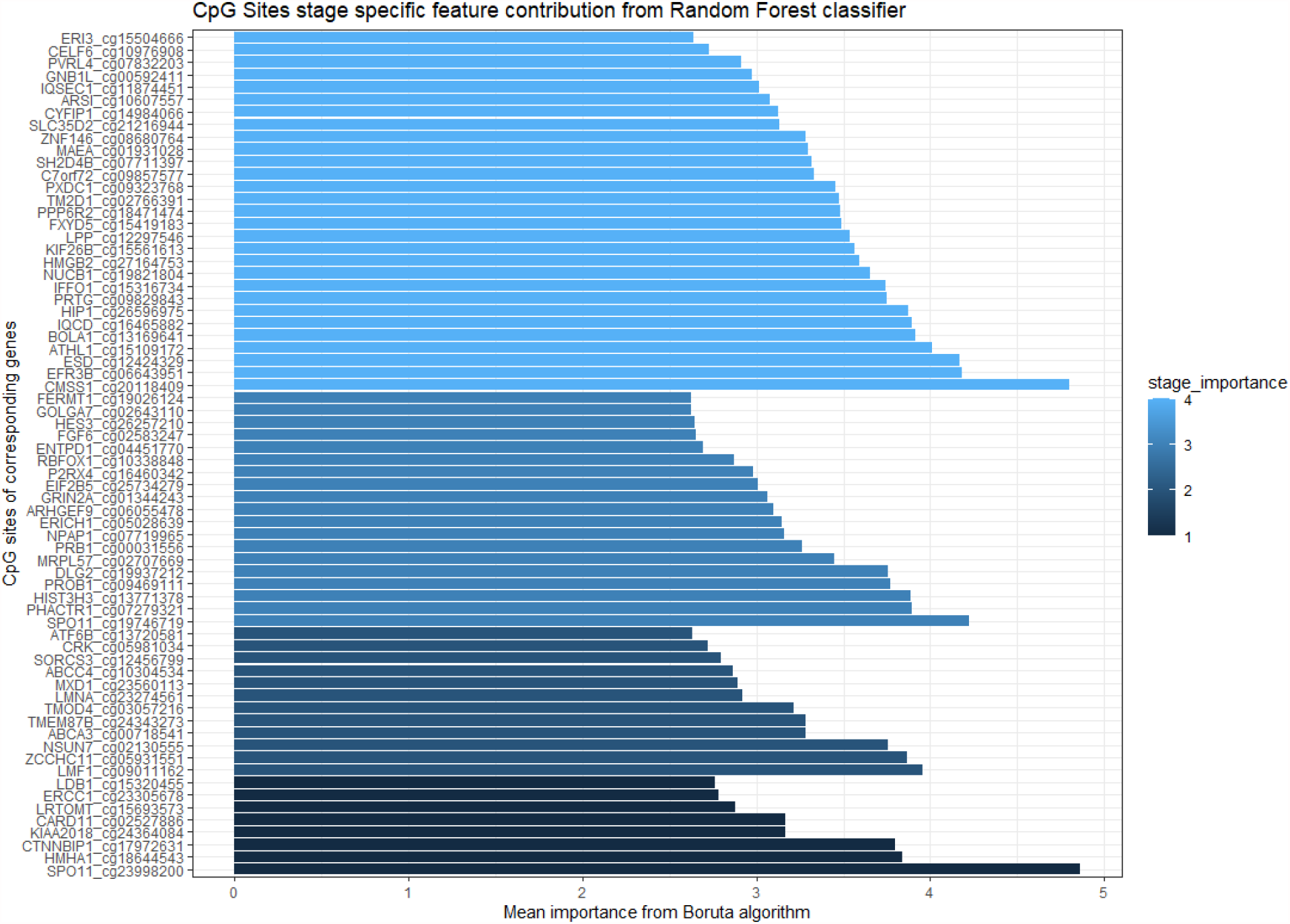
Feature contribution analysis. Feature contribution plot for the pathological stage outcome. X-axis presents the mean importance from the Boruta algorithm where feature contribution for each class (stage) is represented as gradient blue from stage 1 (darker) towards stage 4). From this plot, stage specificity of the CpG sites and their respective genes is assumed based on variable importance from Random Forest classifier.

### 18 genes connect densely in the protein interaction network

After aiming at 67 highly significant genes, the Protein-Protein Interaction (PPI) network of 67 genes corresponding to 68 CpG sites obtained through the Boruta algorithm in Cytoscape 3.8.2 but no interactions were found for PROB1, HES3, FXYD5, and SH2D4B in the human PPI of BioGrid database [28]. Using a Network Analyzer in Cytoscape, we found 2566 nodes and 3374 edges in the PPI (Table 2). Where the top 3 hub genes were LMNA, HIST3H3, and CRK with a high degree of interactions (Figure 4) (Table 3). In the large network of 63 genes, densely connected regions did appear that may represent molecular complexes. To detect densely connected regions, we utilized the MCODE plugin in Cytoscape [29]. We found 1 cluster of 18 densely connected genes with 21 interactions in our PPI network of 63 genes as shown in (Figure 5). Further statistics of network analyzer and MCODE are available in supporting information S1.

**Table 2:**
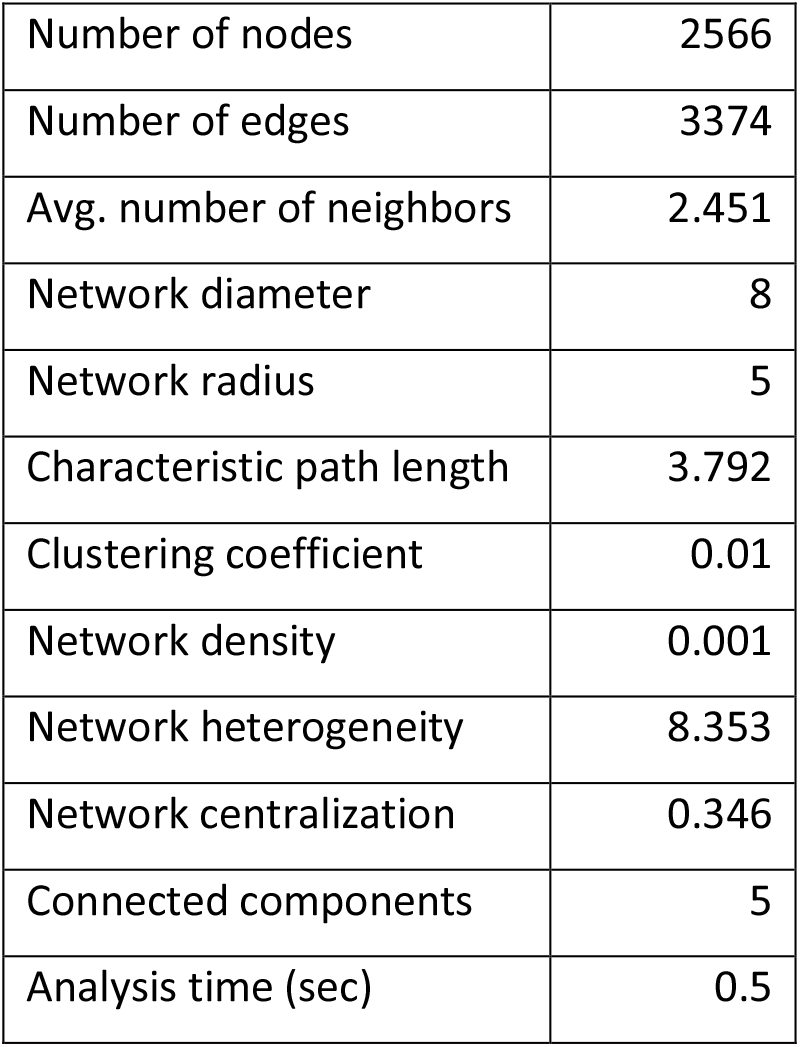
Network analyzer. Summary statistics from a network analysis of 67 genes

**Table 3:** Hub genes. Top 20 hub genes in the interaction network of 67 genes

| name | Degree |
| --- | --- |
| LMNA | 933 |
| HIST3H3 | 396 |
| CRK | 327 |
| CYFIP1 | 99 |
| HMGB2 | 84 |
| PPP6R2 | 81 |
| RBFOX1 | 75 |
| MAEA | 70 |
| ZCCHC11 | 66 |
| LPP | 65 |
| EIF2B5 | 65 |
| LDB1 | 64 |
| HIP1 | 63 |
| ERCC1 | 63 |
| MRPL57 | 55 |
| CARD11 | 47 |
| MXD1 | 42 |
| NUCB1 | 40 |
| DLG2 | 39 |
| ESD | 37 |

**Figure 4:**
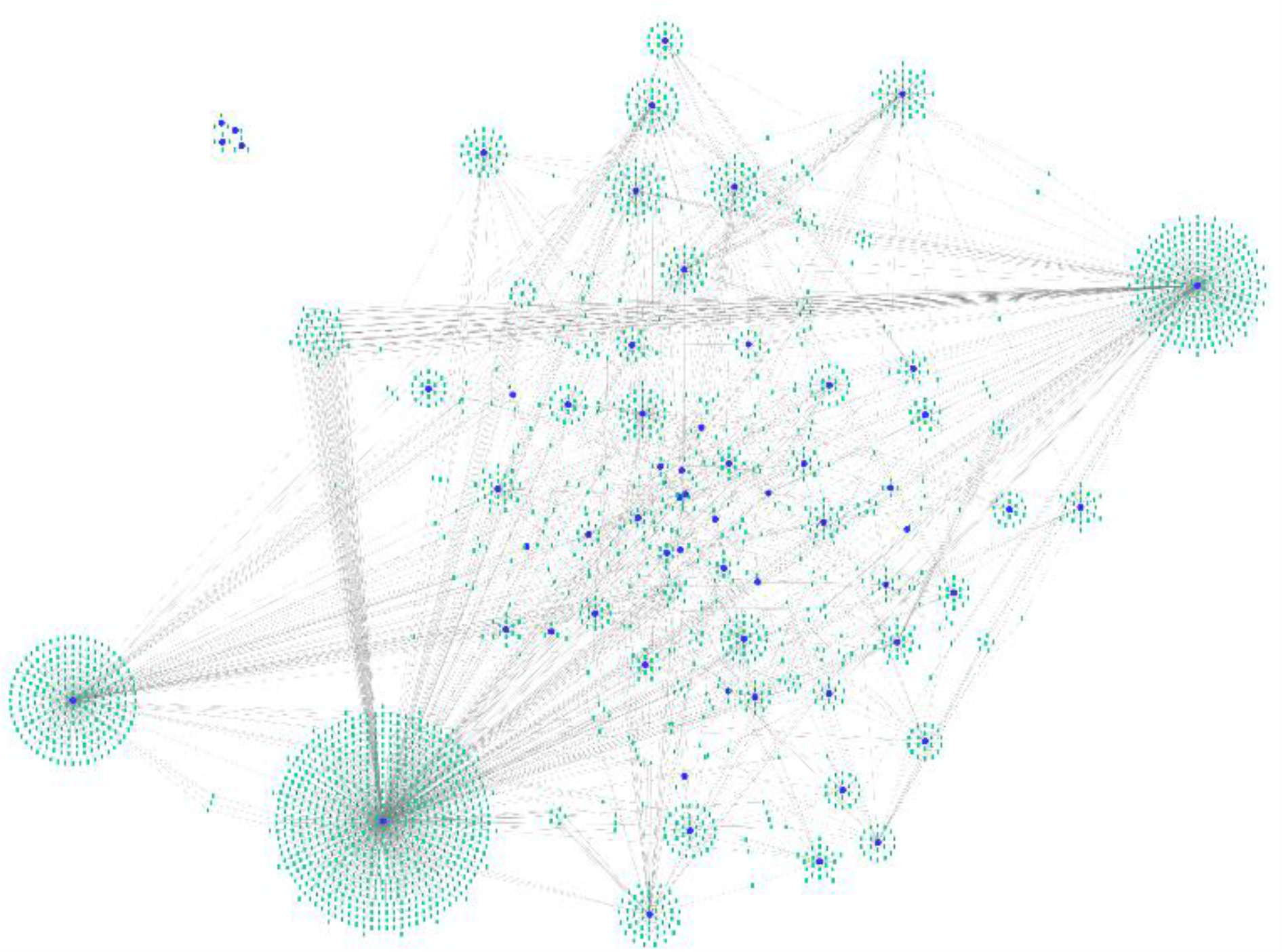
Protein protein interaction network. Protein-Protein Interaction network of proteins encoded by 63 genes with 2566 nodes and 3374 edges where LMNA, HIST3H3, and CRK are identified as top three hub genes.

**Figure 5:**
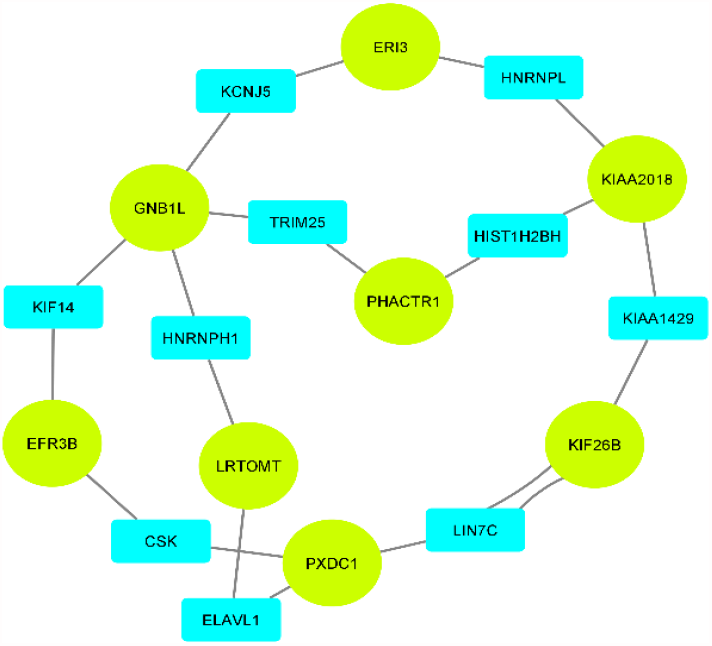
MCODE analysis. Using MCODE, 18 densely connected genes in the interaction network of 63 genes were identified, where ellipses represent the genes.

### Enrichment analysis of genes with high confidence CpG sites

Enrichment analysis was performed for Gene Ontology (GO) and Kyoto Encyclopedia of Genes and Genomes (KEGG) Pathway terms using the EnrichR to further study the 67 genes [30]. The analysis was categorized into KEGG Pathways, Biological Processes, Molecular Function, and Cellular Component with the top terms ranked by adjusted P-value as shown in Figure 6. KEGG and GO enrichment analysis showed that the 67 genes are not significantly related on a functional level. Most genes were found to be encoding enzymes including kinases, ion, and nucleic acid binding proteins which are involved in anatomical structural development, cell differentiation, and proliferation, signal transduction, metabolic and biosynthetic processes. KEGG Pathway analysis revealed that FGF6, CRK, GRIN2A, P2RX4, ABCC4, ATF6B, LMNA, and CTNNBIP1 are involved in cancer-related pathways like MAPK signaling pathway, PI3K-Akt signaling pathway, Wnt signaling pathway, apoptosis, cAMP signaling pathway, and estrogen signaling pathway. A complete list of GO and KEGG terms is provided in supporting information S2.

**Figure 6:**
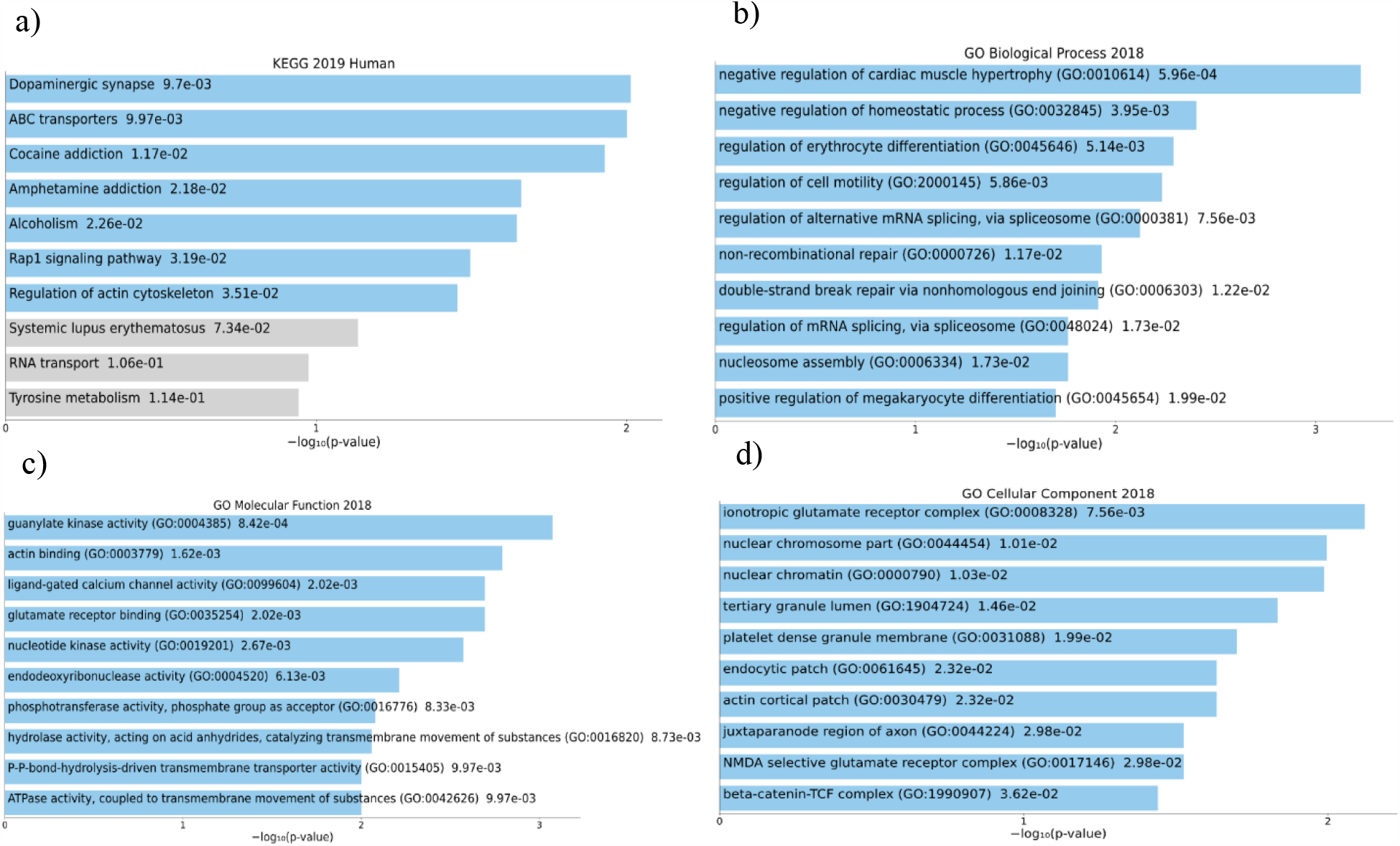
Gene ontology and pathway enrichment analysis. Gene ontology and pathway enrichment analysis of the genes associated high powered CpG sites in HNSCC. (a) KEGG Pathways (b) Biological processes (c) Molecular function (d) Cellular components. The vertical axis represents top 10 GO terms and KEGG pathways while the hortizontal axis represents the statistical significance of the annotations in terms of –log10 (p value).

### Genetic mutations identified in the candidate 67 genes for HNSCC

We also compared our list of prioritized genes with those reported in The Catalogue Of Somatic Mutations In Cancer (COSMIC) and cBioPortal for HNSCC. COSMIC is a comprehensive database of somatic mutations in cancer while cBioPortal contains a wide variety of genomic data which is obtained from multiple cancer types [31]. We explored these databases to identify mutations and copy number alterations in the respective genes for HNSCC. In COSMIC, CARD11, HIP1, LPP are found to be annotated as oncogenes while GRIN2A and LMNA play their role as a tumor suppressor and fusion gene in cancer. Gene mutations including missense, truncating, or inframe mutations were not found in CTNNBIP1, TM2D1, HES3, GOLGA7, EFR3B, and C7orf72 (SPATA48), but there are some copy number alterations in these genes including deep deletions and amplification in HNSCC patients.

## Discussion

Epigenetics, more specifically DNA methylation, appears to play a critical role in cancer progression [13]. HNSCC is the sixth leading cancer type and on the top in the South Asian countries including Pakistan and India, making it a cancer type of interest. In our study, we demonstrated the capability of the Boruta algorithm, a powerful feature selection algorithm, using large-scale DNA methylation datasets from TCGA. The algorithm eliminated 99.98% of irrelevant CpG sites and extracted 68 high confidence CpG sites to classify pathological stages of HNSCC by building three popular machine learning classifiers i.e. Random Forest, Support Vector Machines, and K-Nearest Neighbors. Random Forest was the best classifier where it classified the data points with an accuracy of 90.62%. We evaluated the models by considering kappa statistics and P-Value. Kappa statistics are useful in analyzing the performance of the machine learning classifiers by measuring the match of the predicted labels by the machine learning classifier to the actual labels as a reference [32]. The kappa value for the Random forest was 0.875 while the kappa value for the KNN classifier was the lowest at 0.417. This means the Random Forest is a strong classifier in terms of predicting the pathologic stages of HNSCC where the data (DNA methylation data) used for building this model is 64-81% reliable while KNN is a weak classifier where the data is only 15-35% reliable for this model [33]. On the other hand, the P-Value provides the statistical significance that how well the models have performed in terms of accuracy than the No Information Rate (P-Value [Acc > NIR]), where for all the models the No Information Rate was 25%. These results indicate with the inline hypothesis that if we use the Boruta algorithm before prediction using Random Forest classifier we can predict the pathological stages of HNSCC with significant accuracy.

In previous studies, multiple parameters and different types of integrated data have been used to predict stages of different cancer types using machine learning with good accuracy [34-37], but there was little use of methylation data to classify pathological stages especially in head and neck cancer.

After identifying the set of 68 high confidence CpG sites representing 67 genes with a putative role in HNSCC, we analyzed the feature contribution of the random forest classifier with the available literature to determine the stage specificity of the genes in HNSCC or any other cancer type. In our literature review, we could not found relevant studies related to 32 genes out of 67. In our Random Forest model, 8 CpG sites had high predictive power for class “stage1” but no significant literature was found to support early-stage specificity. However, KIAA2018 (USF3) may be involved in the predisposition of thyroid cancer [38]. Additionally, LDB1 [39], HMHA-1 [40], and CTNNBIP1 [41] are associated with cancer progression and metastasis in HNSCC, melanoma, and thyroid cancer respectively. For class “stage2”, 12 CpG sites contributed the most, in which ABCC4, LMNA, MXD1, ABCA3, and CRK have significant behavior in the early stages of cancer. ABCC4 is highly expressed in lung cancer cell lines to promote cell growth, making it a potential target for lung cancer therapy [42]. LMNA is associated with the regulation of gene expression, cell proliferation, and apoptosis, where its loss of expression is incremental towards advanced stages of breast cancer [43]. MXD1 is thought to be a tumor suppressor and its underexpression can lead to tumorigenesis [44]. ABCA3, its downregulation is associated with early stages of cancer [45] where CRK expression is a prognostic factor of oral cancer and can be a potential target for cancer therapy [46]. However, ZCCHC11 is not specific to the early stage but is associated with metastasis and can also be a therapeutic target [47]. Further, 19 CpG sites had high prediction power for class “stage3” significantly. ENTPD1 (CD39) [48], EIF2B5 [49], and HES3 [50] were related to advance stage cancer while FERMT1 (Kindlin-1) [51], RBFOX1 [52], GRIN2A [53], DLG2 [54] are not found to be stage-specific but have a role in tumorigenesis where no relevant literature was found for 12 out of 19 genes. However, inhibitors of CD39 are now entering clinical trials for cancer therapy [55]. Lastly, 29 CpG sites contributed to class “stage4a”, LPP [56], HMGB2 [57], HIP1 [58], FXYD5 [59], PVRL4 [60], CYFIP1 [61], IQSEC1 (GEP100) [62], and KIF2B6 [63] are involved in different types of cancers related to advance stage metastasis. Additionally, literature stating advance stage relation with NUCB1 was not found, however, NUCB2 enhances cell migration and invasion in the colon and other cancers [64] which is in high similarity with NUCB1 [65]. Moreover, ESD [66], ZNF146 [67], IFFO1 [68], MAEA [69], CELF6 [70], SLC3D2 [71], PPP6R2 [72] are not stage-specific but they can contribute to tumorigenesis. Further, the top gene, CMSS1 (C3orf26), its CpG site ranked highest by the Boruta Algorithm, is not characterized well, however, it can be associated with the immortalization of cancer cells [73]. The feature contribution of a highly significant classification model can be useful for assuming the stage specificity of the genes or their CpG sites, supported by a literature review. In summary, roughly 52% of the genes are involved in tumorigenesis and metastasis, and the remaining 33 key genes make for interesting candidates for in vivo and in vitro analysis for their role in cancer initiation and progression.

In our study, there are three main limitations. First, we were limited to the sample size of 27 cases per stage since we required a balanced set of labels for each pathological stage to avoid class bias; because the maximum number of cases available for stage 1 were 27 while cases for stage 2, stage 3 and stage 4a were over 70. The performance and accuracy of our machine learning models could improve significantly if larger balanced datasets were available. However, the problem of missing information is a trend across datasets [74], which is counteracted by under sampling and oversampling techniques accompanied by information loss and overfitting [75]. Second, we have considered the head and neck as a singular region by itself while in reality, its subtypes including oral, larynx, salivary glands cancer are different and a sufficient amount of data should be available for each subtype. Third, in our dataset, 84 % of the patients come from a European background and only 10 % of the patients come from Africa and Asia while 6 % were not reported for their race. Studies have shown that DNA methylation patterns are known to be different across populations [76, 77]. The datasets should have comprised diverse demographics, which means our results cannot be extrapolated to other populations due to the majority representation of the white patients.

In conclusion, our study identified 68 key CpG sites as potential features using the Boruta algorithm and a Random Forest classifier can be used to predict the pathological stages of HNSCC with significant accuracy. These results demonstrate the promising capabilities of the Boruta algorithm on large datasets like DNA methylation profiles to extract useful insights from the noise. An integrated omics analysis of these findings will help us narrow down the list further of ideal candidates and then the role of those candidates will be needed to be experimentally validated using *in vitro* or *in vivo* models.

## Experimental procedures

### Data collection

We identified and gathered patient data of HNSCC with DNA methylation profiles and confirmed pathological stages in TCGA. DNA methylation data files from TCGA consist of 10 columns while clinical files of the corresponding patients consist of 181 columns or fields. We extracted the Composite element REF, Beta_value, Gene_symbol fields from DNA methylation profiles and Case ID, ajcc_pathologic_stage from clinical files of 529 patients. We found a total of 452 out of 529 primary tumor methylation profiles with confirmed pathological stages that were included in this study (Figure 7).

**Figure 7:**
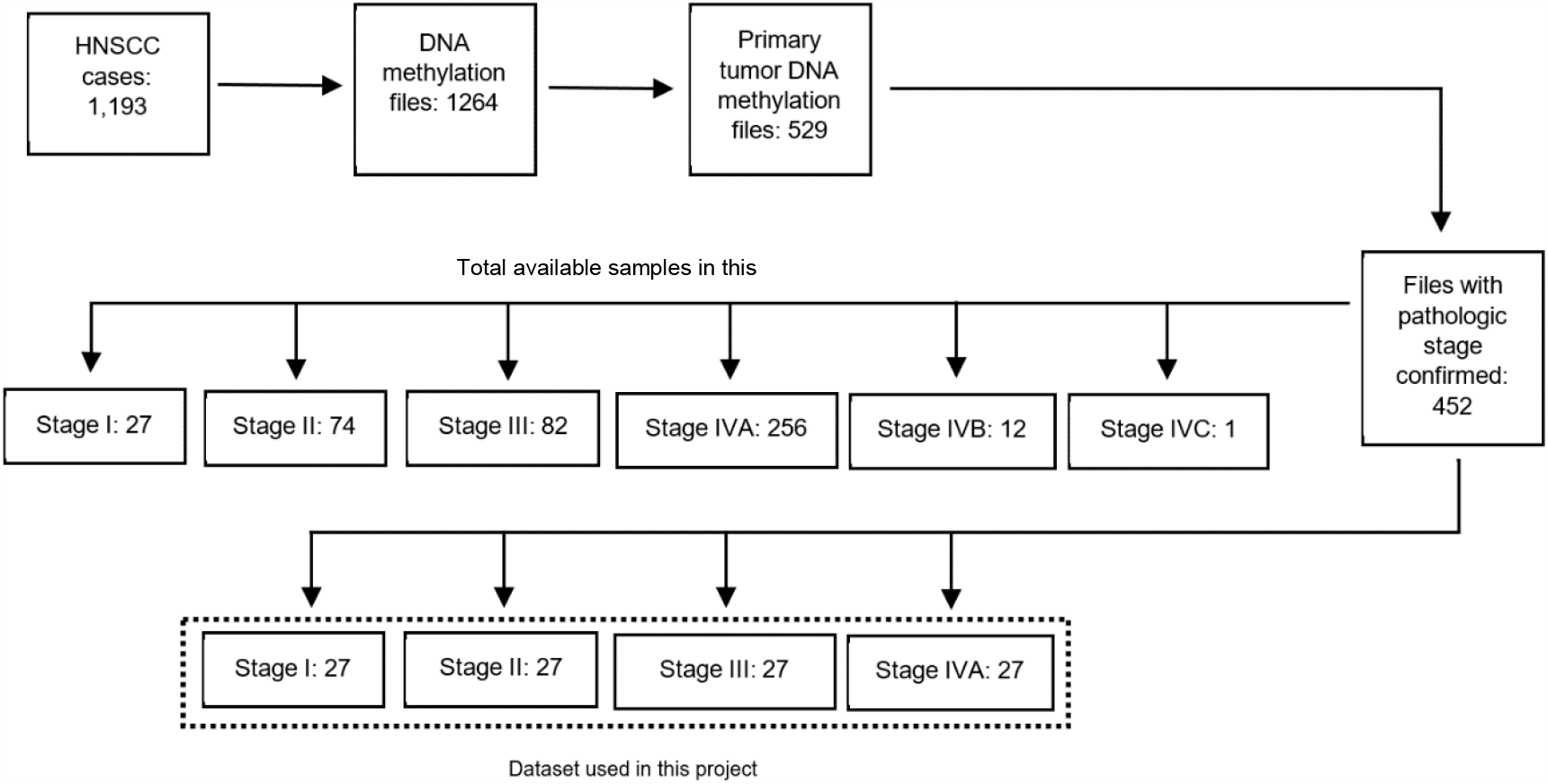
Data availability. There are 1264 DNA profiles from 1,193 HNSCC cases, out of which only 529 DNA methylation profiles for primary tumor samples are available. 452 DNA methylation profiles of 452 primary tumor samples have confirmed pathological stage assigned. DNA methylation profiles available for each stage are not available in equal proportion. A set of 108 DNA methylation files (in equal proportions) of stage 1, stage 2, stage 3, and stage 4a regardless of primary sites is used for training our machine learning classifiers.

### Data pre-processing

We used R programming 4.0.3 and SPSS 22 to pre-process data for the machine learning algorithms. We randomly picked data files of a total of 108 patients of stage 1, stage 2, stage 3, and stage 4a (with 27 from each stage, limited by stage 1 which only had 27 cases) for the training and testing of our classifiers. The data was compiled into the main file which consisted of gene_symbol, composite element REF (CpG sites), and columns of 108 patients consisting of beta values.

### HNSCC mutated genes

TCGA has reported simple somatic mutations in 20,759 genes in HNSCC, out of which we were able to find 354,434 composite element REF (rows) representing methylation/ CpG sites of 18,509 genes, which are then extracted from the main file by joining gene symbols (Figure 8)

**Figure 8:**
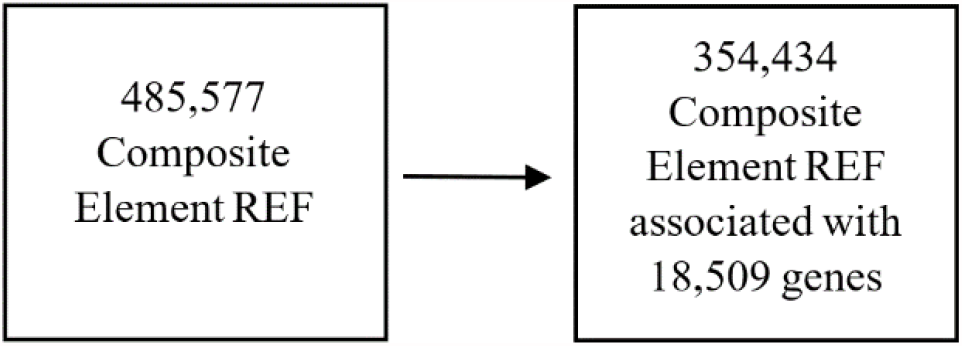
Gene specific CpG sites. Human Methylation 450K BeadChip array containing 485577 probes (CpG sites) covering 99 percent of RefSeq genes. CpG sites in our study are reduced to 294,603 by involving only those CpG sites which are associated with genes that are found to be mutated in HNSCC in TCGA i.e. 20,684 genes.

### Missing Value imputation

Machine learning classifiers do not perform optimally with missing values. Therefore, CpG sites with missing Beta Values for different samples were removed. The composite element REF (CpG sites) was reduced from 354,434 to 294,603 rows (CpG sites).

### Main file transpose

The composite element REF (CpG sites) were used as features (columns), and beta values and stage labels were used as observations (rows). We divided the main file into 148 sub-files and each file was transposed. Each file was repaired after transpose for the feature selection algorithm (Figure 9).

**Figure 9:**
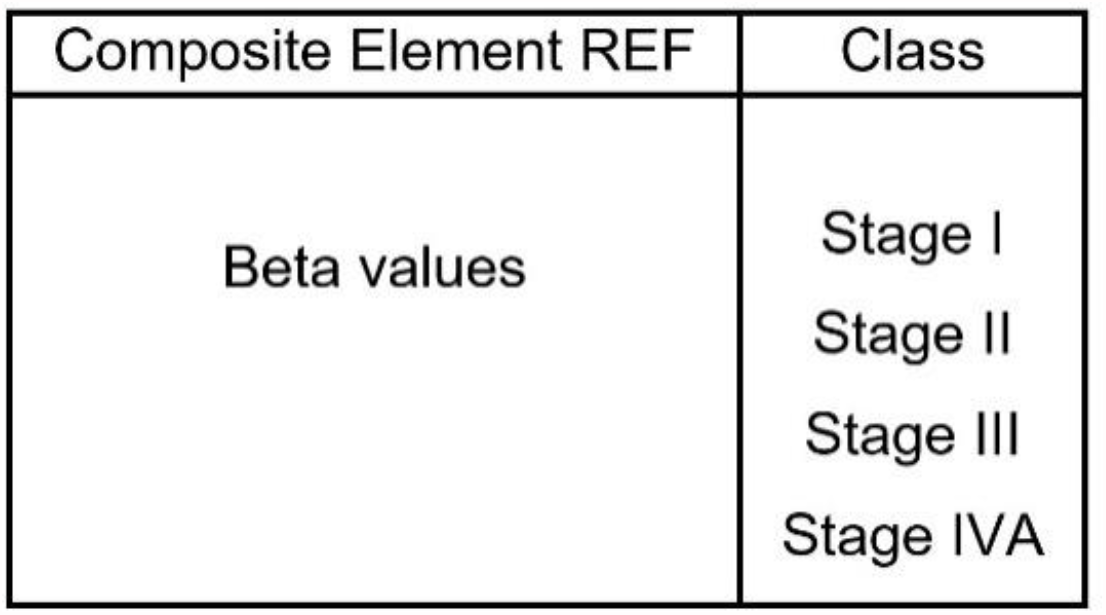
Data frame structure. The data frame consists of 108 rows of Beta values for each Composite Element REF (CpG Site) column. Each row has a specified stage of a particular patient which is placed in the Class column as a response object for the feature selection and machine learning algorithms.

### Feature selection

To identify key CpG sites, we used the Boruta algorithm (package v7.0) [1]. This feature selection algorithm iteratively removes the features (CpG sites) which are proved by a statistical test to be less relevant. This removal of features is done by calculating the maximum Z score among shadow features (shuffled copy of original features to remove correlation) and classifying the features into important and unimportant, where the unimportant features are removed [1]. The algorithm was performed in 3 phases in R 4.0.3 with default parameters, however, max runs for phase 1 were set to 20,000 while max runs for phase 2 and 3 were set to 50,000. The CpG sites obtained in phase 3 are used as features in the machine learning algorithms to classify pathological stages of HNSCC (Figure 10).

**Figure 10:**
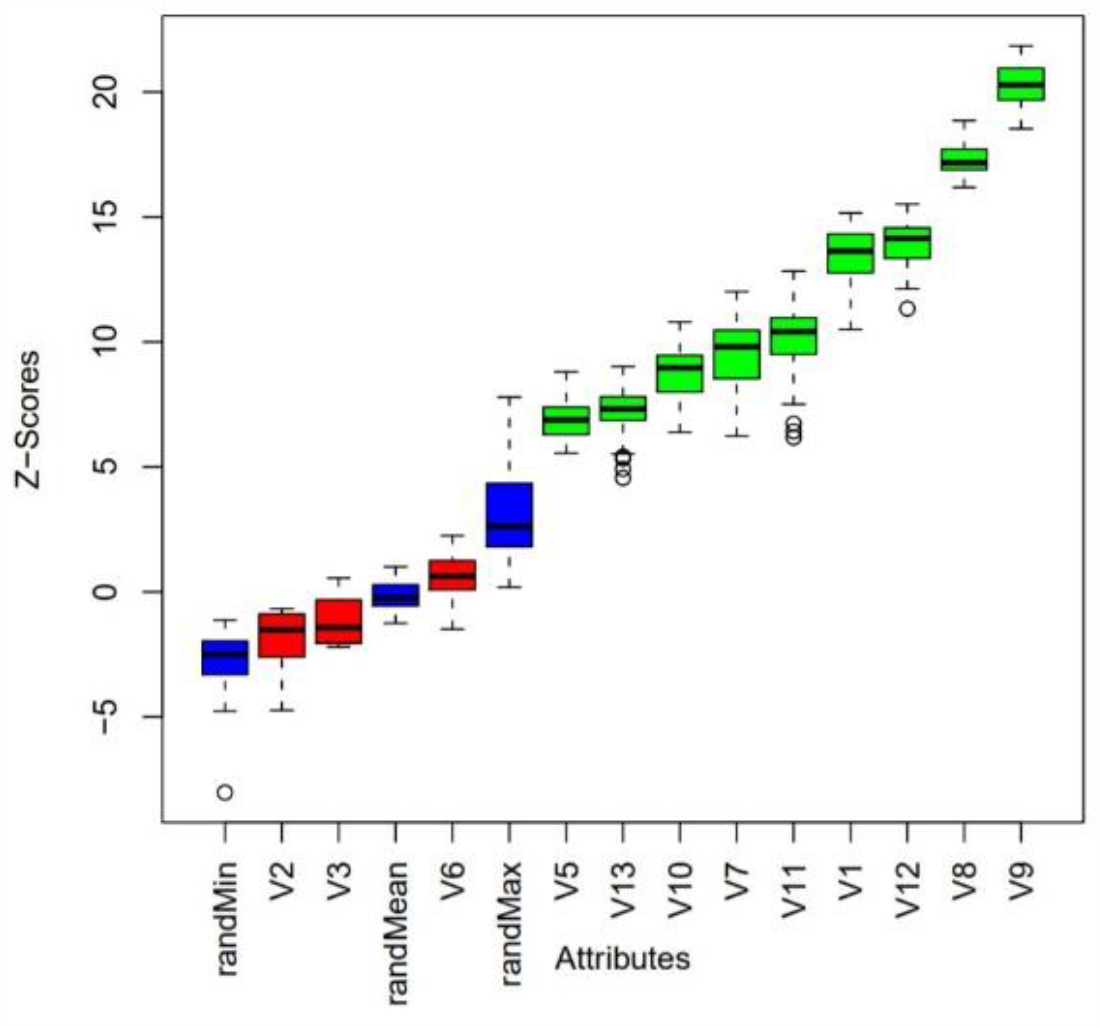
Boruta result plot. Result plot of Boruta algorithm performed on example ozone data adapted from Kursa *et al* (2010). Blue boxplots corresponds to minimal, average and maximum Z score of shadow features which are the randomized copy of original features. Red and green boxplots represent Z scores of respectively rejected and confirmed features [1].

### Machine learning models

We developed three different machine learning models using the random forest, support vector machine (both linear and radial kernel), and k-nearest neighbor using the caret package in R [78]. Validation rule was set to 30 times repeated 10 fold cross-validation using trainControl function of the caret package while methods in the train function used for random forest, support vector machine with linear kernel, support vector machine with radial, and k-nearest neighbor were “rf”, “svmLinear”, “svmRadial” and “knn” respectively. For the evaluation of machine learning models, confusion matrixes were built to compare accuracies, Kappa Values, and P-Values of the models. All the algorithms are performed on an Intel machine of 8th Gen Core i7 with 1.8 GHz base speed and 16 GB Ram.

### Network and enrichment analysis

For network analysis, the protein-protein interaction (PPI) network of proteins encoded by genes associated with the prioritized CpG sites was constructed using Cytoscape version 3.8.2. The source of the network interaction database was the Biological General Repository for Interaction Datasets (BioGIRD release 4.2.191, human taxon identifier: 9606). The Network Analyzer (version 4.4.6) was used to compute the network summary statistics. Moreover, Molecular Complex Detection (MCODE) was used to identify densely connected regions of genes in the PPI network. “Degree cutoff = 25”, “node score cutoff = 0.2”, “k-core = 2”, and “max. depth from seed = 100” was set as the cut-off parameters. Enrichment analysis of GO annotation was performed using EnrichR (https://maayanlab.cloud/Enrichr/).

Additionally, (Figure 11) illustrates the overall methodology followed in this research.

**Figure 11:**
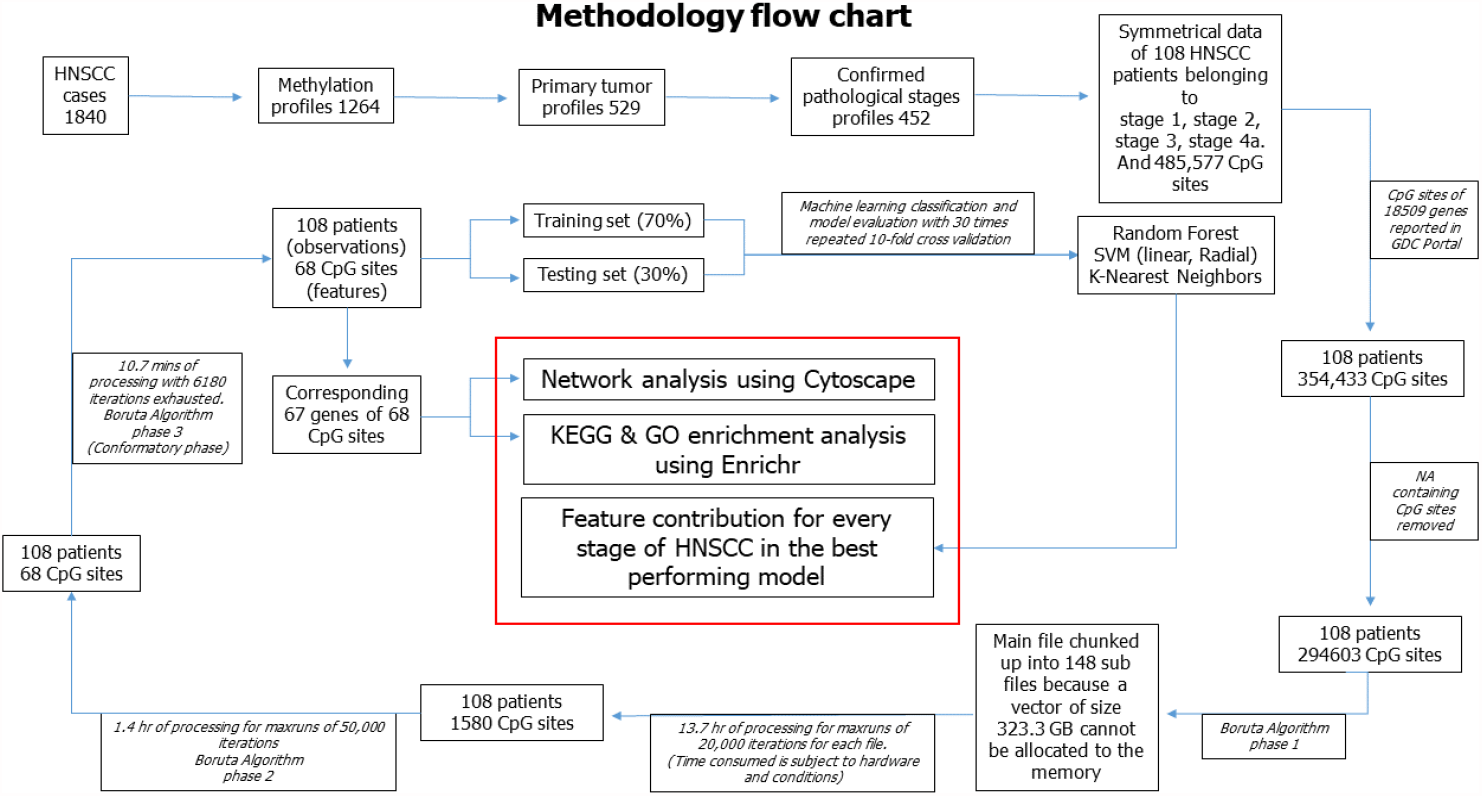
Research methodology. TCGA contained 1264 methylation profiles for all 1840 head and neck cancer patients out of which only 529 primary tumor files were available. We selected the datasets of patients with their comfirmed pathological stages. For the machine learning models, a symmetrical dataset was compiled of 108 HNSCC patients with confirmed pathological stages and methylation profiles. The methylation profiles consist of more than 485K CpG sites which were reduced to 384K CpG sites associated with 18509 genes. The CpG sites are aggressively reduced to 68 in 3 phases using the Boruta algorithm and based on these 68 CpG sites, machine learning models with 10 fold Cross-Validation repeated 30 times and 70% training split. Finally, network and enrichment analysis was performed for the 67 genes corresponding to 68 CpG sites and feature contribution was also analyzed for all four stages. Feature contribution provides the measure of influence of features (CpG sites in our case) on the prediction outcomes [19], where the outcome built were the pathological stage of HNSCC.

## Data Availability

All the data is publicly available at The Cancer Genome Atlas (TCGA). Case IDs of randomly selected patients is available in the supporting information S3. The R code for building machine learning models is available at Arsalan_Riaz/Rcode_patterns.zip at https://github.com/PML-research/Arsalan_Riaz

## Data and code availability

All the data is publicly available at The Cancer Genome Atlas (TCGA). Case ID’s of randomly selected patients is available in the supporting information S3. The R code for building machine learning models is available at “Arsalan_Riaz/Rcode_patterns.zip at main · PML-research/Arsalan_Riaz · GitHub“.

## Acknowledgments

This project would not have been possible without the generous support from all members of the Precision Medicine Lab (PML), especially Shahram Khan and Shagufta Rehmat. We are grateful to the Higher Education Commission (HEC) of Pakistan who through their National Center for Big Data and Cloud Computing (NCBC) have provided funding and support for this project. We would also like to show our gratitude to CECOS University and Rehman Medical Institute (RMI) for being partners in this unique academia-industry complex at the Precision Medicine Lab (PML).

## Author Contributions

Conceptualization, methodology and writing – original draft, A.R., F.K.; network and enrichment analysis, M.S.; discussion on study design, S.Z., A.S.; writing – review, editing and supervision, F.K.

## Supporting information

Supporting files are available at “Arsalan_Riaz/Supporting_information at main · PML-research/Arsalan_Riaz · GitHub “

S1: Network and MCODE analysis

S2: GO and KEGG terms of 67 genes

S3: Case ID’s of samples

